# Non-Adherence of PLWHA in taking Antiretroviral during the COVID-19 pandemic in West Sumatra, Indonesia: Qualitative analysis

**DOI:** 10.1101/2022.06.26.22276906

**Authors:** Adriani Suwito, Ikhwana Elfitri, Elly Usman, Arina Widya, Evi Hasnita, Ria Ningsih

**Affiliations:** Public Health departement, Andalas University, West Sumatra-Indonesia; Faculty of Engineering Andalas University; Faculty of Pharmacy, Andalas University; Medical science Andalas University; Stikes Fort De Kock Bukitinggi; M.Jamil Hospital Andalas Padang (head of VCT service) Indonesia

**Keywords:** Non-adherence, PLWHA, Covid-19, HIV/AIDS

## Abstract

**Introduction:** COVID-19 pandemic condition affects the adherence of PLWHA in taking antiretroviral drugs. People with Lost to Follow-up (LTFU) were an average of 26 %, The feared impact is the high mortality rate and decreased productivity, visits of PLWHA patients to M Jamil Hospital come from all regions in West Sumatra to the southern provinces of Jambi and Bengkulu. This study aimed to determine factors that influence patient non-Adherence during the COVID-19 pandemic.

**Method:** This study was conducted by exploring the experiences of patients and health workers by observing adherence to taking antiretroviral, 34 informant: 25 patients, 5 NGOs, and 4 health workers, The data were collected from September 6^th^ to October 9^th^ 2021, Analysis data using N VIVO 12.

**Results:** Patients in taking medication during the COVID-19 pandemic were feeling bored, most of the patient lived was so far, so patients often asked for help from NGOs to take medication. Forgetfulness, busyness, and fear of being exposed to COVID-19. Then, the economic factor, due to the large number of layoffs, and the absence of BPJS insurance affects compliance, while the factors of stigma, stress, feeling well are slightly expressed by patients. It turns out that the Covid-19 condition affects patients to come for treatment to the hospital. In addition to drug side effects, dizziness, nausea, and vomiting also affect medication adherence. The role of NGOs in providing information and education as well as motivating and taking medication during the COVID-19 condition is very important.

**Conclusion:** The most common causes of non-compliance from loss to follow-up during the COVID-19 pandemic are; bored, far from home, forgetful, and economic factors for fear of covid-19. Most of the side effects of drugs are dizziness, and nausea and vomiting so that the role of NGOs is important in helping patients take medication.

**Strength and limitation of this study**

➢ This qualitative research is about medication adherence during the covid-19
➢ The factors that influence patient adherence to taking ART are different during this covid-19 pandemic, namely the distance from which they live has an effect on patients wanting to take medication, so the role of NGOs in sending drugs is very important, according to the doctors approval, after the next 3 months the patient is required to come, this is never happened before the Covid-19 condition, Patients adherence are very dependent on the patient’s self-awareness to live in the long term, the convenience has been provided by M Jamil Hospital by making it easy to send drugs to the patient’s own address.
➢ This qualitative analysis using N vivo 12, which has now developed into N vivo 14
➢ Patient recruitment is divided into two, namely obedient and non-compliant patients, but the patient’s answers are almost the same.

## A1. INTRODUCTION

Covid-19 pandemic increases the threat of death in society. It can also attack the immune system which can cause death.

People with HIV/AIDS has dysfunction of the immune system, which increases the mortality rate(1). PLWHA is a person with HIV / AIDS who is infected with a virus, destroys the immune system and destroys CD4 cells(2)AIDS is a collection of symptoms of reduced self-defense ability caused by the entry of HIV into a person’s body that attacks the body’s system.(3),(4)So that people with HIV/AIDS can live a long time if they adhere to antiretroviral therapy (ART). If they do not comply, the death will be accelerated, and the amount of virus in the blood or viral load cannot be suppressed, or treatment failure will occur.(5).Non-adherence results in drug resistance; resistance will decrease, and the disease will worsen and can cause death(6)The number of people with HIV/AIDS in Indonesia until March 2021 was 543,100 people. The rate of non-compliance or loss to follow-up/LFU was 26%, while the allowable standard was below 20%, the mortality rate for AIDS sufferers was relatively high in 2021, namely 61,192 people (11.2%) compared to 262,693 people (48%) who have received therapy antiretroviral(7).

West Sumatra is one of the provinces in Indonesia which is famous for its strong culture and religion. The cumulative number of HIV/AIDS cases until 2021 in West Sumatra was 4,484 people(8)The non-compliance rate for HIV/AIDS cases for treatment at M. Jamil Hospital Padang was 30% during this COVID-19 pandemic. The results of the interview with the head of the VCT out service room, it is stated that the most common causes of patients being absent during the COVID-19 pandemic, such as patients not coming because they could not afford insurance (BPJS). Another reason is because many have been laid off, besides being afraid to take medicine due to Covid-19. Based on the above phenomenon, researchers are interested to know the causes of non-adherence to taking antiretroviral drugs in PLWHA.

## 2. METHOD

### 2.1 Study design

This study used a qualitative design. It focused on the factors related to non-adherence to HIV/AIDS patients in taking medication during the COVID-19 pandemic. The number of primary informants was 25 people, with criteria for HIV patients who had received ART therapy for at least three months, cooperative patients, and not in an advanced stage. Then, 5 supporting informants were added, namely PLWHA who accompanied the Taratak Foundation, West Sumatra on September-December 2021, 4 health workers, namely; doctors, head of poly VCT people, pharmacy staff. With purposive sampling technique.

### 2.2 Recruitment participant

Recruitment of informants was carried out between September 6th to October 9th 2021. This research was conducted M. Jamil Hospital Padang. This hospital is a referral from 19 districts and cities, sufficient to meet the area of West Sumatra. These goals were related to the causes of non-adherence to taking ART drugs, among others; forgetting, stigma, tired of taking medicine, busy at work, far from home, side effects of drugs, mentoring or assistance, family support, fear of covid-19. Participants were followed up for up to 4 months.

### 2.3 Interviewer

Interviewers have been trained to collect socio demographic data (age, identity, etc.). The interview process was recorded by a tape recorder. The interview guide format is different for doctors, staff, assistants, and HIV/AIDS patients. Documentation and field observations were also carried out. When conducting interviews with patients, appointments were made. The main informants were patients who did not comply with the treatment program, were still alive, and had stopped ART. The question items contain the factors that cause non-adherence to HIV/AIDS patients using ART during the COVID-19 pandemic.

### 2.4. Patient and Public involvement

The research questions was developed started since the covid-19 pandemic, there are many patient has lost to follow up ART (26%), many efforts have been made by nurse and even by NGOs to reduce that, for example by calling patients, visit to the patient’s home but they are not very meaningful. This is causes of how we are interested to knowing the reason why patients non-adherence. A part from that, the experience of patients and NGOs also mentioned that covid-19 pandemic make a lot of problems affecting patient in taking drugs. This support the research question; what are the factors that influence PLWHA non-adherence, how to overcome the problem of non-adherence to HIV/AIDS patient taking antiretroviral therapy (ART).

Patient involve in this study were selected randomly and grouped between obedient and non-adherence, the patient’s expression and complaints were almost the same since the pandemic covid-19. The number of patients interviewed until they were saturated.

In the recruitment process, patient were not involved, but NGOs were involved, who were responsible for several patient and they understood which patient were adherence or non-adherence, patients gave the information about how to take medication and consequences if they are not discipline to take ART.

The result of this study are expected to be very useful for patient, where the factors cause of non-adherence can be known, education is easier to provide, NGOs will easier to motivate patients to take medication discipline. Disseminating the results of this research can be through education presentations, and writing to this journal.

The burden of the intervention was assessed by the patient through their treatment discipline, the also seen from the medical record administration, and the evaluated by NGOs.

### 2.5. Data analysis

Descriptive data were analyzed to describe the characteristics of people living with HIV/AIDS(9) by using N Vivo 12.

### 2.6 Ethical Statement

This research had been obtained permission from the Ethics Committee of. M.Jamil Hospital Padang No. 363/KEPK/2021. All study participants provided written informed consent and had an understanding of the study. Informed consent was given to informants at the time of the patient face it, There is no conflict of interest here in this study.

## 3. RESULT

### 3.1 Characteristics of participants

Qualitative research was conducted from 6 September to 9 October 2021. Qualitative data were obtained through in-depth interviews with 34 informants, with details of 25 HIV/AIDS patients, four officers, and five assistants/NGOs. The data can be described in the following table 1:

**Table. 1.** Characteristics of in-depth Interview Informants.

| Characteristics | f | % |
| --- | --- | --- |
| **Age** |  |  |
| 15-19 years old | 0 | 0 |
| 20-24 years old | 4 | 12 |
| 25-49 year | 29 | 85 |
| >50 years | 1 | 3 |
| **Gender** |  |  |
| Man | 29 | 85 |
| Woman | 5 | 15 |
| **Profession** |  |  |
| Health worker | 4 | 12 |
| NGO | 5 | 15 |
| businessman | 10 | 29 |
| trade | 4 | 12 |
| government | 2 | 6 |
| employees | 1 | 3 |
| housewife | 4 | 12 |
| Company /PT | 3 | 9 |
| College student | 1 | 3 |
| agriculture | 3 | 9 |
| **Education** |  |  |
| JUNIOR HIGH | 11 | 32 |
| SCHOOL | 3 | 9 |
| Senior/vocational | 9 | 26 |
| high school | 8 | 24 |
| Students | 0 | 0 |
| D3 |  |  |
| S1 |  |  |
|  | 34 people | 100% |

Table 1 shows that 85% of the age group is 25-49 years old, they are of productive age, 85% of them are male, most of their occupations are 29% self-employed, and 32% are informants with high school education.

### 3.1 Causes of Patient Non-Adherence

From 25 patients interviewed, 13 patients stated that they forgot to take their medicine and 12 patients never forgot to take their medicine, according to the patient’s statement as follows;

“I never took medicine because I forgot (informant (inf) 14, inf 12, inf 23, inf 29)”, “I never took medicine one month ago (inf 16), “I never took medicine one year ago ((inf 19)”, “I have stopped taking medicine for up to 7 years (inf 18)”, “Did you ever take medicine when you were young (inf 20)”, “I often go one month without taking medicine (inf 21)”, “Once, the referral time ran out (Inf22)”, “Once, I was late for taking my medicine for eight months (Inf 28), “Once, I dropped my medication because I returned to another city (Inf 34).”

Figure 3.1.1 describes the causes of non-adherence to PLWHA patients in West Sumatra by using N Vivo 12:

**Figure 3.1.1.**
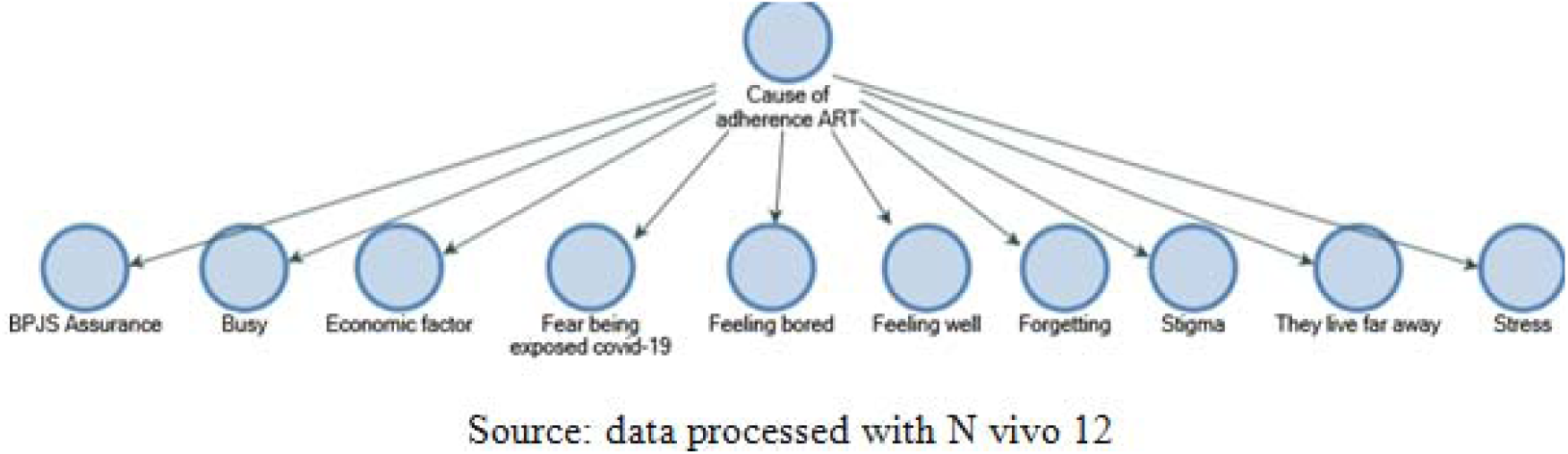
Overview of causes of non-Adherence to patients living with HIV in taking ART.

The general description of the causes of non-adherence to take medication for PLWHA patients taking HIV antiretroviral during the COVID-19 pandemic that the patient expressed, among others, is as follows; afraid of covid-19, then because of stress, there was a stigma in society towards HIV patients, the reason being busy at work was also the cause of forgetting to take medicine, feeling bored or bored with taking medicine for life, the distance from where you live from home to take medicine to the hospital too affect, because economic factors have a huge impact on society because during the Covid-19 pandemic many suffered as a result of dropping out of work (PHK), so they were unable to pay BPJS insurance contributions for treatment, then also conveyed by patients even by VCT Poli doctors that often patients feel they have recovered or feel fine.

From the description of the causes of non-compliance that were mostly expressed by patients during the COVID-19 pandemic, it can be seen in Figure 3.1.2 below that the data was processed with N VIVO 12.

**Figure 3.1.2.**
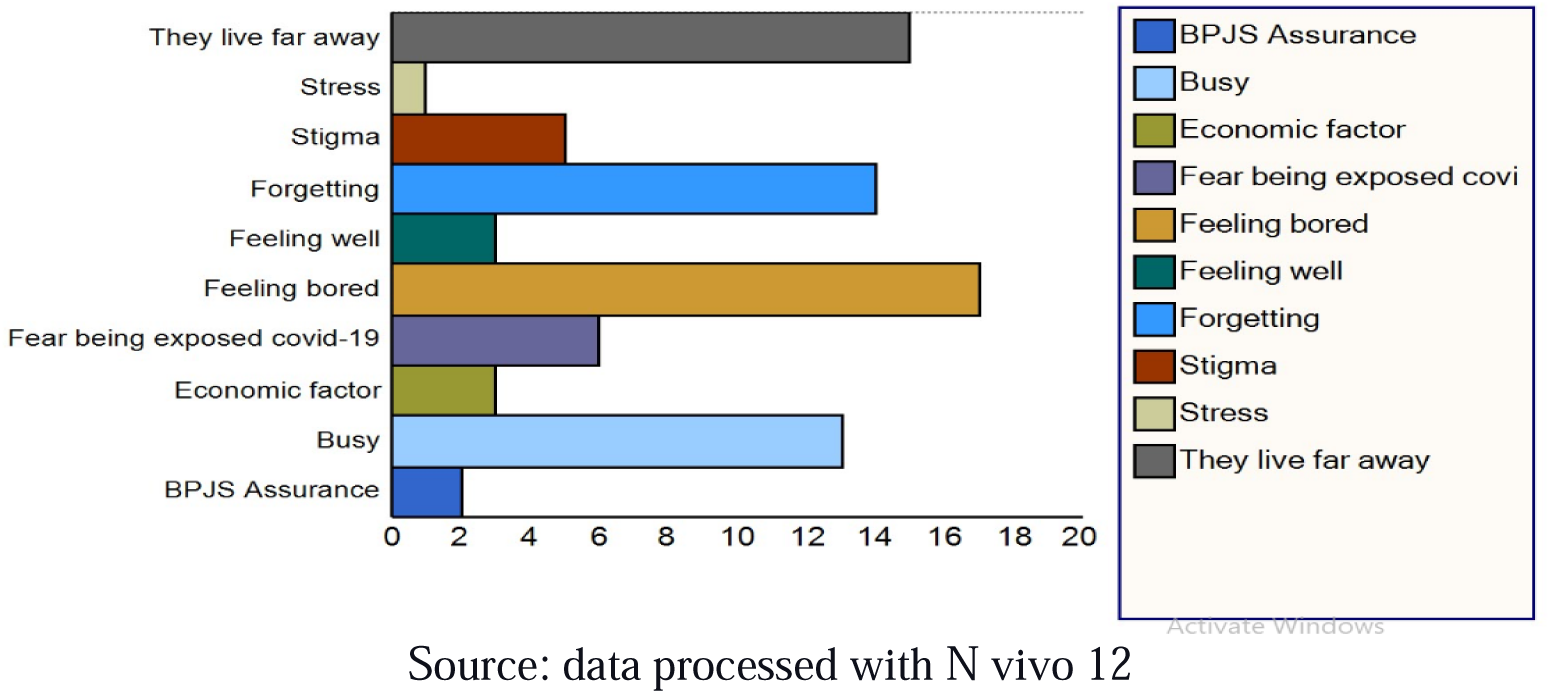
Dominant description of the causes of non-Adherence in PLWHA patients using ART.

The results of interviews with researchers found that the five factors that caused non-adherence to PLWHA patients using ART during the COVID-19 pandemic were mostly due to; Feeling bored and tired of taking medicine because they have to take it for life, secondly because they lived far away, so patients often asked for help from NGOs to pick up medicine and deliver it to the patient’s house. Patients came from all provinces in West Sumatra. The farthest were South Beach and Darmasraya, with a travel time of 5-6 hours. Some even came from outside the region, such as Jambi and southern Bengkulu. The delivery of drugs during the COVID-19 pandemic was allowed after obtaining permission from the doctor and the Head of the VCT Hospital Polyclinic. M. Jamil Padang. Referring to the Regulation of the Director General of P2P Ministry of Health No; 060/SPK-1/MPS/2022.(10). The next cause was forgetting; It was related to the busyness of PLWHA patients in their daily activities. The fourth reason was the economic factor due to the many layoffs (PHK) that occurred during the COVID-19 pandemic, which dramatically affected the patient’s economy. In contrast, 29% of the jobs were self-employed, which was directly impacted by the pandemic conditions. The fifth dominant cause was the fear of being exposed to COVID-19— generally patients who are afraid of being exposed to the Covid-19 virus.

### 3.2 Side effects of antiretroviral drugs

The next cause of non-adherence was due to drug side effects. From the results of interviews with patients and doctors, as well as VCT polyclinic officers, a general description of drug side effects can be seen in Figure 3.2.1.

**Figure 3.2.1.**
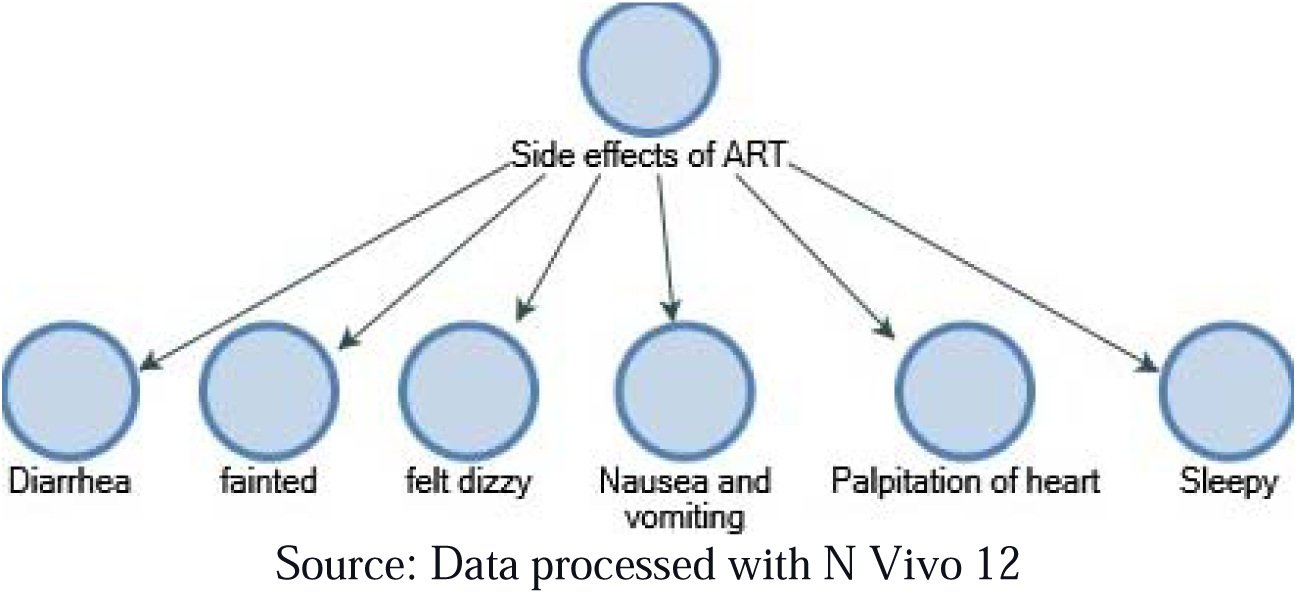
Overview of side effects of antiretroviral drugs in PLWHA.

The side effects are different for each patient, so one patient said that if he took the medicine he felt dizzy, so he often stopped treatment. Others said that they fainted or fell after taking medication, then complained of being often sleepy, complaints of nausea and vomiting being the most common. Then diarrhea or diarrhea was often complained of by the patient, the last complaint felt by the patient was heart palpitations. The third most dominant side effects of using antiretroviral drugs in PLWHA can be seen in Figure 3.2.2, namely dizziness expressed by 18 patients, nausea and vomiting expressed by 8 patients, while side effects of drugs that caused drowsiness were expressed by 2 patients, complaints This side effect is not the only one expressed by the patient.

**Figure 3.2.2.**
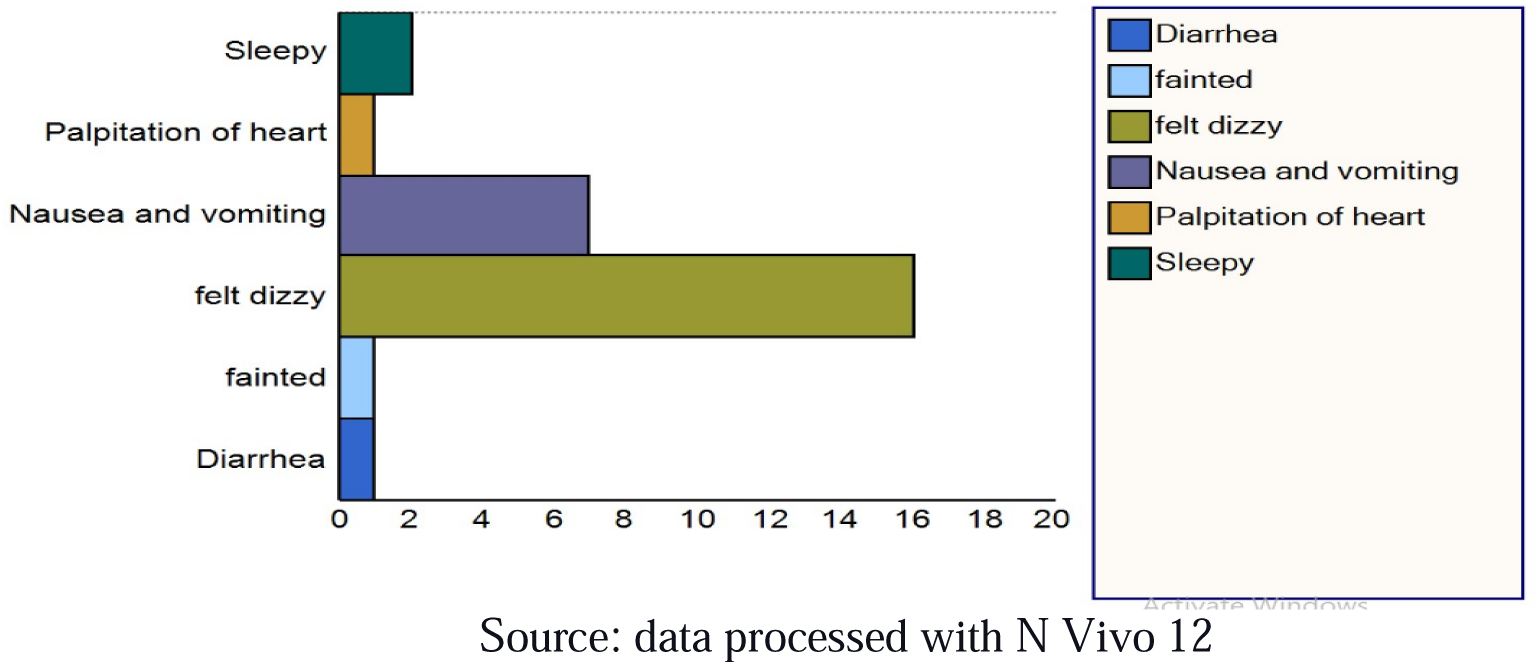
Description of the most side effects in PLWHA.

Then for other symptoms such as diarrhea, heart palpitations, feeling hot, and fainting, from each of them only one person complained of such conditions. This side effect needs to be paid attention to by health workers, both doctors and nurses because this can be the cause of patients not continuing their treatment.

### 3.3. The role of NGOs in helping PLWHA patients

The results of the research on the role of NGOs during the COVID-19 pandemic are shown in Figure 3.3.1 below; 25 patients revealed that 9 patients said that the role of NGOs mostly was to provide educational information, then 6 patients said that the role of NGOs was to motivate patients to be disciplined in taking medicine, after that it was revealed by 6 other people that NGOs also helped get drugs, while 4 people don’t want to be associated with NGOs. All these NGOs are present at the hospital every day, their activities are monitoring the patient’s discipline to come for treatment and discipline to take medicine. This is done in collaboration with the head of the outpatient polyclinic and the outpatient poly doctor. During the Covid-19 condition, the role of NGOs is very important.

**Figure 3.3.1.**
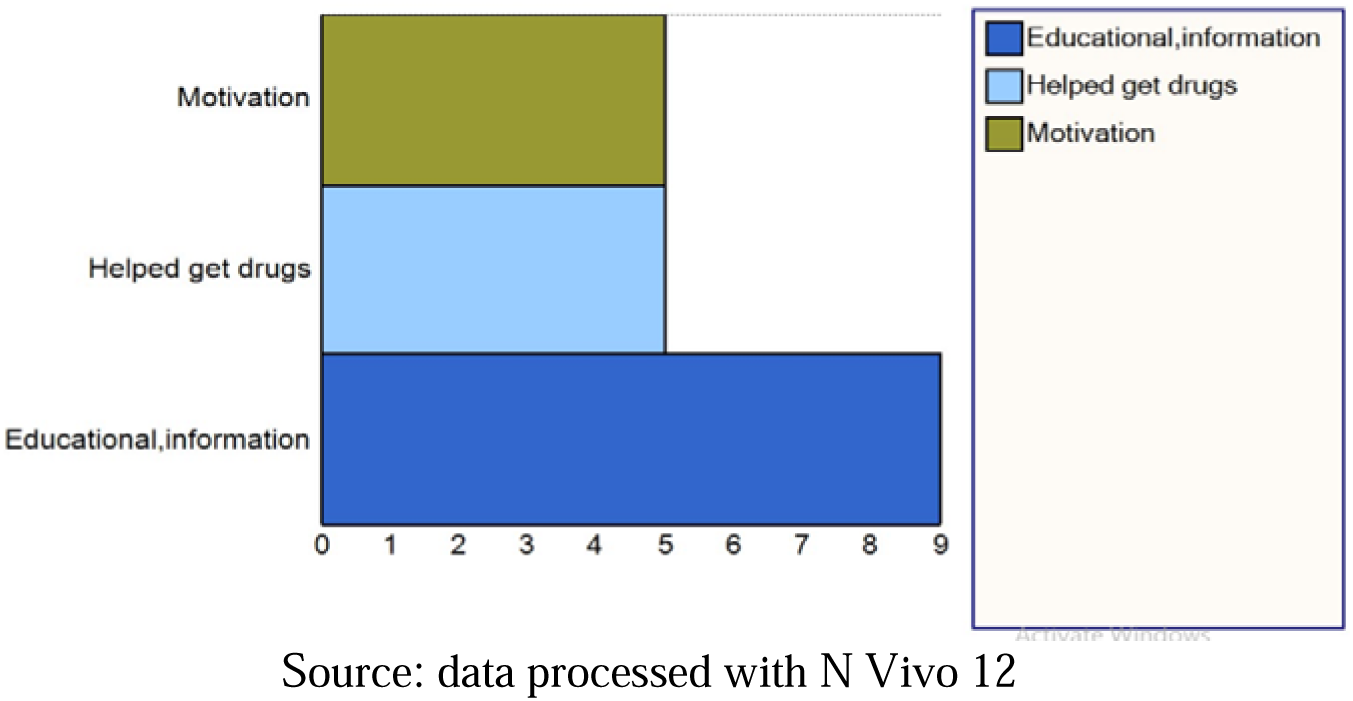
The role of NGOs in helping PLWHA patients.

## 4. DISCUSSION

### 4.1 Causes of non-adherence

During the COVID-19 pandemic conditions, PLWHA patients in West Sumatra had a patient adherence rate of 74% or still quite low, PLWHA can be adherent between 80% and 95% to maintain viral suppression(11). This was shown by the results of a qualitative study which stated that the cause of non-compliance of PLWHA patients taking medication was due to boredom in taking medication, respectively. this is an expression that is often told by patients, this is because PLWHA patients have to take medication for the rest of their life, then the next cause was because the distance from where they live was far from the hospital, the farthest distance was 5-6 hours away, so NGOs are often assisted to taking the patient’s medication. This result is in line with Kalichman’s research(1).The social distancing factor during the COVID-19 pandemic condition influences patients to take medicine. Then forgetfulness was also the cause of patients not taking medication, this is because patients are busy working or doing activities, so PLWHA patients must be reminded by NGOs to come for treatment checks. Then the condition of being afraid of being exposed to COVID-19 also causes patients to not comply with taking medication, this is in line with Hasan Ejaz’s research(12)said that HIV-infected patients are at high risk of getting COVID-19 due to their low immune system. The reasons are, among others, because their immune system is weak, they will easily get sick, besides that other causes are because of the stigma, feeling fine, and the absence of BPJS insurance, so patients cannot take medicine, because the insurance card is not active. again because they are unable to pay the mandatory insurance premium, this is due to economic factors, where many layoffs have occurred, since the Covid-19 condition, the last cause of non-compliance is stress, this happens to patients who just found out the diagnosis. This research different with a systematic review by Heestermans et al, said that the sociodemographic, psychosocial, health status, treatment-related and intervention-related determinants are interlinked and contributed to optimal adherence(13)

Generally, patients tend to hide that they are taking antiretroviral drugs, and do not want to other people know the condition of the disease, These results are in line with research conducted by Eswarachar (14), It is stated that the causes of non-adherence to PLWHA patients taking medication include: age, work status, distance, side effects of drugs, number of pills, education, and mental health of patients, fatigue, being away from home, busy, and forgetful, and the belief that the drug is not available.. The influence of alcohol is also a cause of non-compliance(15)(16). In the research of Andreas et al. in Africa, it was found that 41.8% of LFU patients had 6.0% died within five years of starting ART(9). In line with the research of Kalichman et al in Atlanta, Georgia, in 2020(1), the difficulties of PLWHA patients with the spread of Covid-19 interfere with HIV care. They block access to places of service including to get food(4)

### 4.2 Side effects of Antiretroviral Therapy

One of the reasons for the patient’s lack of adherence to taking medication is that there are side effects of drugs that are quite influential, from the results of the research that were disclosed by the patient and after the data was processed with N vivo12, it was seen that many patients said that the effects of taking medication were dizziness, nausea and vomiting, especially after taking the first line drug, namely Efavirenz, of course, patients who have just received this therapy must really be given good education, so that patients understand the importance of taking medication, and understand that PLWHA patients take the drug for the rest of their life, if there are side effects feelings such as dizziness, nausea and vomiting, drowsiness, palpitations are common symptoms that are felt at the beginning of taking the drug, but the drug should not be stopped, which is why it needs a deep understanding from the patient, different things from the research revealed by Nathan(17)In a systematic review of 21 observational studies of drug use during the COVID-19 pandemic it was found that HIV patients who received Lopinavir/ritonavir is not certain to improve clinical outcomes or prevent infection in patients who are at high risk of contracting COVID-19 and according to WHO, its usefulness will be reviewed. Then, the availability of ART has been effective in turning HIV infection into a chronic disease so that people living with HIV have a life expectancy that is close to normal, in elderly patients due to comorbidity associated with age and the presence of organ dysfunction and physiological changes that are at risk for reactions due to drug use, continuous education is needed. about good patient management(18), especially during the COVID-19 pandemic. So patients who do not experience drug side effects are more likely to adhere to antiretroviral treatment than those who experience drug side effects(19).

### 4.3. NGO’s Role

During the COVID-19 pandemic, the role of NGOs in helping PLWHA patients is very important. Researchers directly observed the activities of NGOs at the VCT Poly, which most often included; strengthening through the provision of information and education so that patients continue to seek treatment and assist all the needs of patients who are their responsibility to obtain drugs. The NGO helps deliver drugs via expedition to the address of the patient requesting assistance. These NGOs are always present at the VCT out service every day at M.Jamil hospital Padang. They are under the coordinator of the Medan Plus foundation, with a contract of work number SK no; 060/SPK-1/MPS/2022, and is financed with the support of the Global Fund Funding program and the salary is determined by the Spritia foundation. In the Covid-19 condition, NGOs can help patients get medicine for patients in accordance with the approval of the doctor and the Head of the VCT outpatient polyclinic.

Patients can take the drug for a 2-3 month supply. The following third month the patient had to return to the hospital to monitor his health. If the patient does not come, the NGO helps to call the patient, asking how the patient’s condition is. If the patient’s condition does not allow to come, the NGO will discuss with the head of the polyclinic what steps will be taken to help the patient. The results of the discussion between the NGO and the Head of the Polyclinic decided that patients should be assisted as soon as possible, they even made home visits to patients who did not come for treatment to prevent LTFU. This is an extraordinary service performed by M. Jamil Hospital Padang. This is in line with the research of Irene At al 2020 in Ghana, which said that during the COVID-19 pandemic,(20)This is also in line with the research of Angela Kaida et al. which states that the role of peer support is to facilitate HIV patients interacting with the community, their contribution and motivation in antiretroviral treatment in PLWHA is very large.(21)(22). the aim of this NGO activity is to help them to get medicine and convince patients to take medicine in their daily life(4)

This is also in line with research in America in 2020 that the role of social workers during the covid-19 pandemic is very important(23). In line with Smith et al said that covid-19 has disrupted routine health service delivery and there is increasing use of digital interventions to reduce exposure of patient and health care workers to Covid-19 infection(24).The facilitator or NGO gave their testimony in motivating him to always be disciplined and take his medicine on time. The benefits of this NGO activity are felt by HIV/AIDS patients who seek treatment at the VCT out service patient M.Jamil hospital Padang. Currently, 5 NGOs are working to help patients seek treatment in VCT outpatient services. The role of NGOs in providing education and information is also carried out for patients who experience fear during the COVID-19 pandemic. Some patients experience a fear of opening up about their status to their families, even shutting down to remain silent and take responsibility for their health. PLWHA often only talk to NGOs(25). This is in line with Adelekan’s research, 2019 which states that stigma and lack of support are the main causes of LTFU(26), the assistance in question is from accompanying the patient in expressing his feelings. Important social systems are built to improve patient survival to continue treatment(27) to reduce the level of morbidity and mortality and reduce the risk of HIV/AIDS transmission, it is very necessary for the long-term suppression of viral load. So that the provision of ART in the long term is expected to achieve stability in the 90-90-90 HIV/AIDS program(28).

## 4. CONCLUSION

The conclusion of a qualitative study on this research; boredom in taking medicine, a place to live far away, especially since the Covid-19 pandemic condition is very influential. So that patients often asked for help from NGOs to deliver drugs to patients’ homes. The next factor is forgetting to take medication, then because of the busy schedule of PLWHA patients. The next reason is because of fear of covid-19, so that PLWHA patients need to ask NGOs for help to get drugs with the doctor’s approval and send them to the patient’s home. Then, economic factors and the absence of insurance affect the patient’s adherence to taking medication, one of the causes that the patient disclosed was the large number of layoffs, the dominant side effects of the drug were dizziness and nausea and vomiting. So the role of NGOs in providing information and education as well as motivating and consuming medicines during the Covid-19 condition is very important. These side effects need to be a concern for health workers, both doctors and nurses, the aims for this condition is patients not adhering to taking medication.

## Data Availability

All data produced in the present work are contained in the manuscript

## AUTHOR’S CONTRIBUTION

AS, IE, EU, AW,EH designed the master study, the AS designed the qualitative and analysis. RN contributed to data collection and coding, interpretation of AS, IE, EU, AW, results and editing of manuscripts. AS contributed to funding, study design, interpretation of results, and editing of manuscripts.

## UCKNOWLEDGEMENTS

The authors would like to thanks the individuals who participated in this study, the team at the outpatient VCT of M. Jamil Hospital Padang-West Sumatra, Indonesia. And for the data collection and coding team, including Raveinal MD, Bude Ns, Hidayat, Miko.

